## Supplementary figures and images for "Spatial analysis of COVID-19 spread in Iran: Insights into geographical and structural transmission determinants at a province level"

### Association between explanatory variables

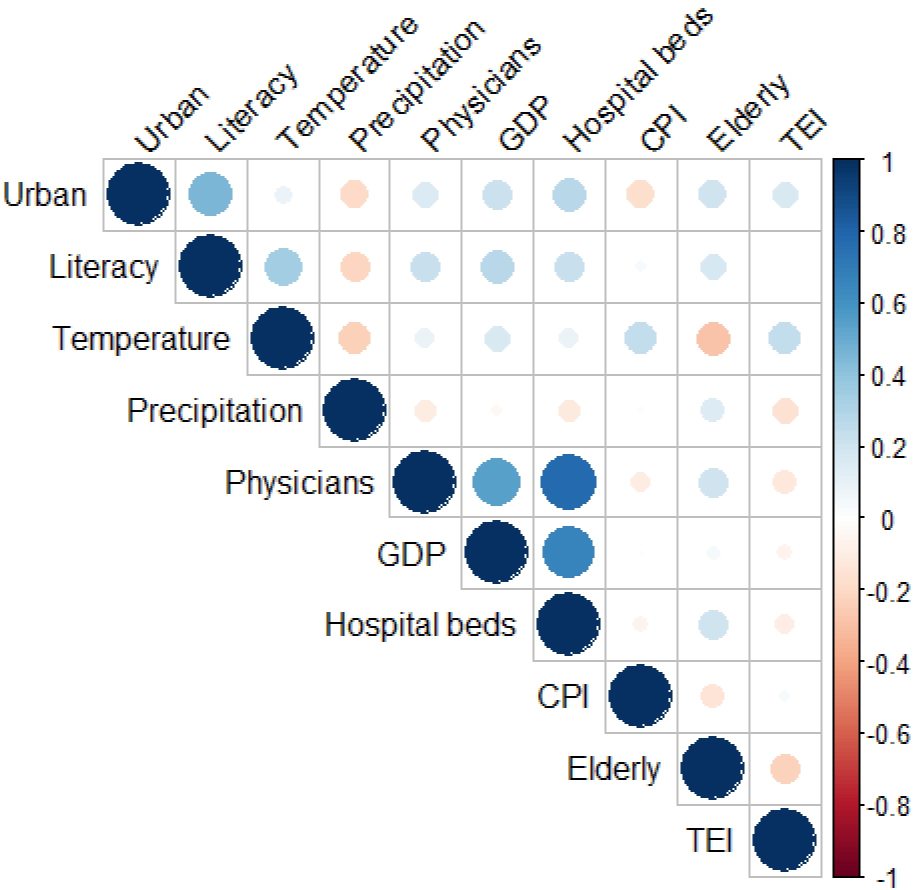
